# Endoscopic submucosal dissection for superficial esophageal squamous cell carcinoma with esophageal gland duct involvement: A single-center study on safety, efficacy, and outcomes in 748 cases

**DOI:** 10.64898/2026.07.26.26358953

**Authors:** Yanan Wang, Zhiwen Li, Hai Wu, Xinlu Zhao, Shangtao Mao, Zhenyu Wang, Ying Yuan, Qiange Ye, Yanmei Zhu, Ying Xiang, Qin Huang, Lei Wang, Guifang Xu

**Author notes:** Corresponding author: Guifang Xu, Tele; Lei Wang; Qin Huang.

## Abstract

**Background:** Endoscopic submucosal dissection (ESD) of superficial esophageal squamous cell carcinoma (SESCC), with or without the esophageal gland duct involvement (DI or NDI), has been reported in small case series, but ESD-related safety, efficacy, and outcomes remain unknown. We conducted a retrospective study of those issues in consecutive 748 patients treated at our center in China.

**Material and Methods:** Over the study period from January 2014 to January 2019, we identified 748 eligible SESCC patients who were divided into the DI (22.6%, 169/748) and NDI (77.4%, 579/748) groups, based on histopathologic reports. Following the European and Japanese guidelines on endoscopic resection of SESCC, we investigated and statistically compared characteristics of clinicopathology and ESD-related safety, efficacy, and prognosis between DI and NDI groups.

**Results:** The mean age of patients was 64.9 years for the cohort and significantly older in the DI (66.1) than in the NDI (64.5) group (P<0.05). There was no significant difference in smoking/alcohol abuse and comorbidity. Endoscopically, the DI group showed significantly larger tumor size and deeper invasion than the NDI group. There was no ESD-related death in the cohort. Although the *en* bloc resection rate was 100%, the complete and curative resection rates were significantly lower in the DI (88.2% and 61.2%, respectively) than in the NDI (96.4%, and 86.9%, respectively) group (P<0.01). ESD-related infection (0.7%) and bleeding (2.3%) rates were very low and no significant difference was found between the two groups. Only one perforation (0.1%) occurred in the NDI group. Post-ESD stenosis was 11.9% for the cohort and significantly more common in the DI (20.7%) than in the NDI (9.3%) group (P<0.01). The overall survival rate was 95.7% for the cohort and significantly lower in the NDI (94.6%) than in the DI (100.0%) group (P<0.05). However, there was no significant difference in disease-specific death, recurrence-free survival, distant metastasis, and the requirement for subsequent chemoradiation therapy and surgery between the two groups.

**Conclusion:** In our cohort, there was no ESD-associated death; ESD-related infection and bleeding were minimal; and perforation was rare. Post-ESD stenosis was 11.9% in prevalence and successfully managed by endoscopic dilation. The overall survival rate was 95.7%. Compared to the NDI group, the DI group showed older age, larger tumor size, deeper invasion, lower complete and curative resection rates, higher prevalence of post-ESD stenosis; there was no significant difference in disease-specific death. Taken together, ESD was safe with excellent efficacy and prognosis for endoscopic resection of SESCC with or without esophageal gland duct involvement.

## Introduction

According to the global cancer statistics of the World Health Organization (WHO) in 2020, esophageal cancer ranks the seventh most common cancer and the sixth commonest cause of cancer-related deaths in the world with more than 604,000 new cases diagnosed and 544,000 deaths every year^1^. The highest incidence of esophageal cancer is in East Asian countries, primarily in China, with heavy socioeconomic burdens^2, 3^. In China, the most common histologic type of esophageal cancer is squamous cell carcinoma with a dismal 5-year survival rate of only 30.3%^4^. At present, the only realistic way to dramatically improve outcomes of these patients is early detection with prompt resection at the disease early stage, such as superficial esophageal squamous cell carcinoma (SESCC), defined as mucosal and submucosal squamous cell carcinomas with or without the presence of lymph node metastasis^5^. It is well known that SESCC frequently involves the ducts of esophageal submucosal glands that are scattered throughout the esophagus^6^. The ducts of these glands are lined with stratified basaloid squamous epithelium with or without a surface columnar cell layer and drain the secretory fluids of submucosal glands into the lumen of the esophagus for lubrication and helping in digestion^6, 7^. In esophageal squamous cell carcinoma, the ductal epithelium may become dysplastic and further progress to carcinoma *in situ* over time before invasive carcinoma is fully developed^7^. Once diagnosed, SESCC may be resected surgically, or preferably endoscopically with endoscopic submucosal dissection (ESD) because of the benefits of minimal injury, but maximal safety and functionality^8–12^. Recently, there have been some studies on endoscopic resection of SESCC with ductal involvement (DI), but the number of patients in those reports is small, and the ESD-related key issues, such as safety, adverse events, and prognosis are rarely discussed^13, 14^. Therefore, the aim of this study was to thoroughly investigate safety, efficacy, and long-term outcomes of ESD in endoscopic resection of SESCC with DI in a large cohort from a high-volume tertiary referral medical center in China.

## Methods

### Patient enrollment

All patients with esophageal lesions, which were resected with ESD over the period from January 1, 2014 through December 31, 2019, were searched in our prospectively established electronic database, regardless of age or sex, stored in the Department of Gastroenterology of the Nanjing Drum Tower Hospital in China. Excluded were patients with multi-focal SESCC, pathologically confirmed non-SESCC lesions, and patients with incomplete clinical or pathological data. As a result, 748 consecutive patients were eligible for this study (Figure 1). These eligible cases were divided into two groups, *ie*, with or without the involvement of the duct of esophageal glands determined histopathologically. Thus, 169 were assigned to the DI group and 579 to the NDI (NDI) group (Figure 1). We reviewed medical history, hospital course, results of laboratory tests, chest and abdominal computerized tomography, electrocardiogram, and preoperative endoscopic examination, such as white-light upper endoscopy, magnified narrow-band imaging, Lugol chromoendoscopy, and endoscopic ultrasonography in all study patients. We recorded the location, and macroscopic type of SESCC, and the endoscopic mucosal “intrapapillary capillary loop” (IPCL) type of each SESCC tumor^15^. All patients were informed of ESD-related risks and signed the ESD procedure consent form. Their private information was deleted in the dataset to protect their privacy. The Medical Ethics Committee of the Nanjing Drum Tower Hospital approved this retrospective study protocol (The approval number: 2023-137-01).

**Figure 1.**
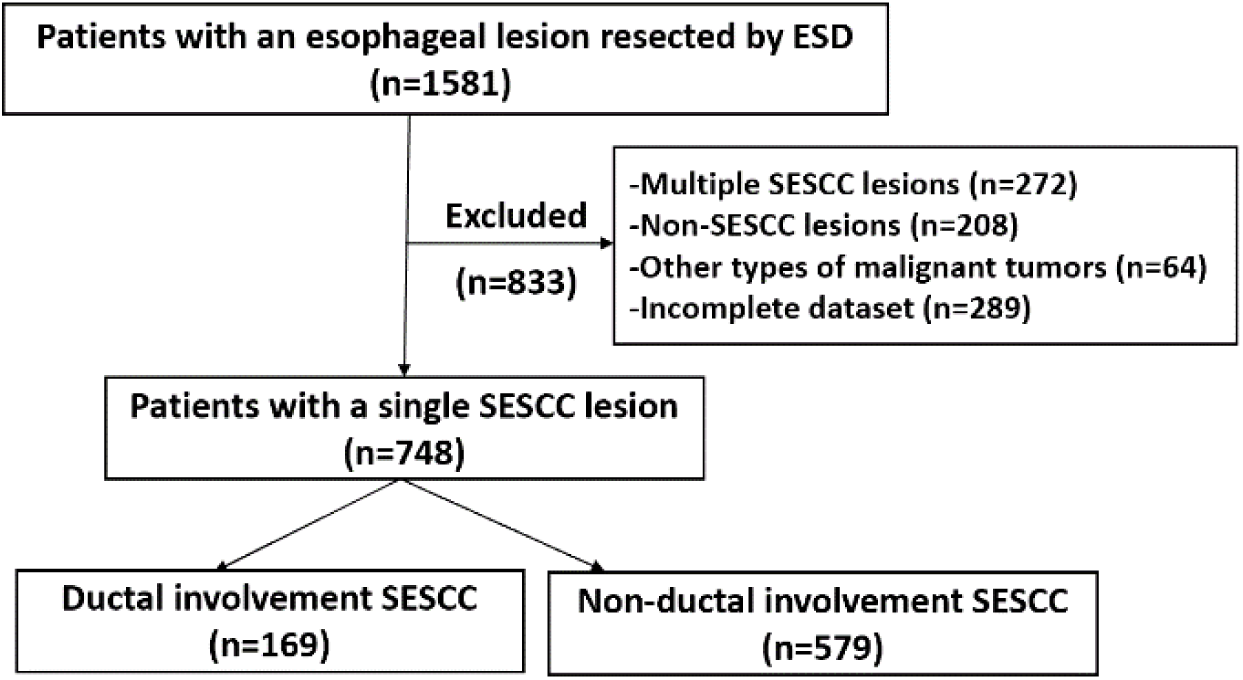
Flow diagram for patient selection and grouping. ESD: endoscopic submucosal dissection; SESCC: superficial esophageal squamous cell carcinoma; n: number.

### ESD procedure

After confirmation of the absence of deeper lesions in the submucosa, nodal and distant metastases with endoscopic ultrasonography and computerized tomography, a routine ESD procedure was performed with an upper endoscope (GIFQ260J; Olympus Corp. Japan), equipped with a single accessory channel for a water jet system and a transparent cap attached to the tip of an endoscope. Before the start of ESD, chromoendoscopy was carried out after directly instilling Lugol’s solution (1.5%) through the endoscope’s biopsy channel to better map the margin of a lesion. With the Dual knife (Olympus Corp. Japan) or T-type ESD knife (MICRO-TECH, Nanjing, China), fresh mucosal cautery dots were made 3-5 mm away from the lesion edge to delineate the perimeter of a mucosal tumor. After an injection of a mixture of normal saline and methylene blue with diluted epinephrine (1:100,000) into the submucosal layer around the tumor to lift the lesion, a circular incision of the mucosal lesion was made and the submucosal layer was dissected with a Dual or T-type knife. All visible active bleeding vessels on the resection wound bed were immediately coagulated by hot biopsy forceps (MICRO-TECH, Nanjing, China) to prevent delayed bleeding (Figure 2).

**Figure 2.**
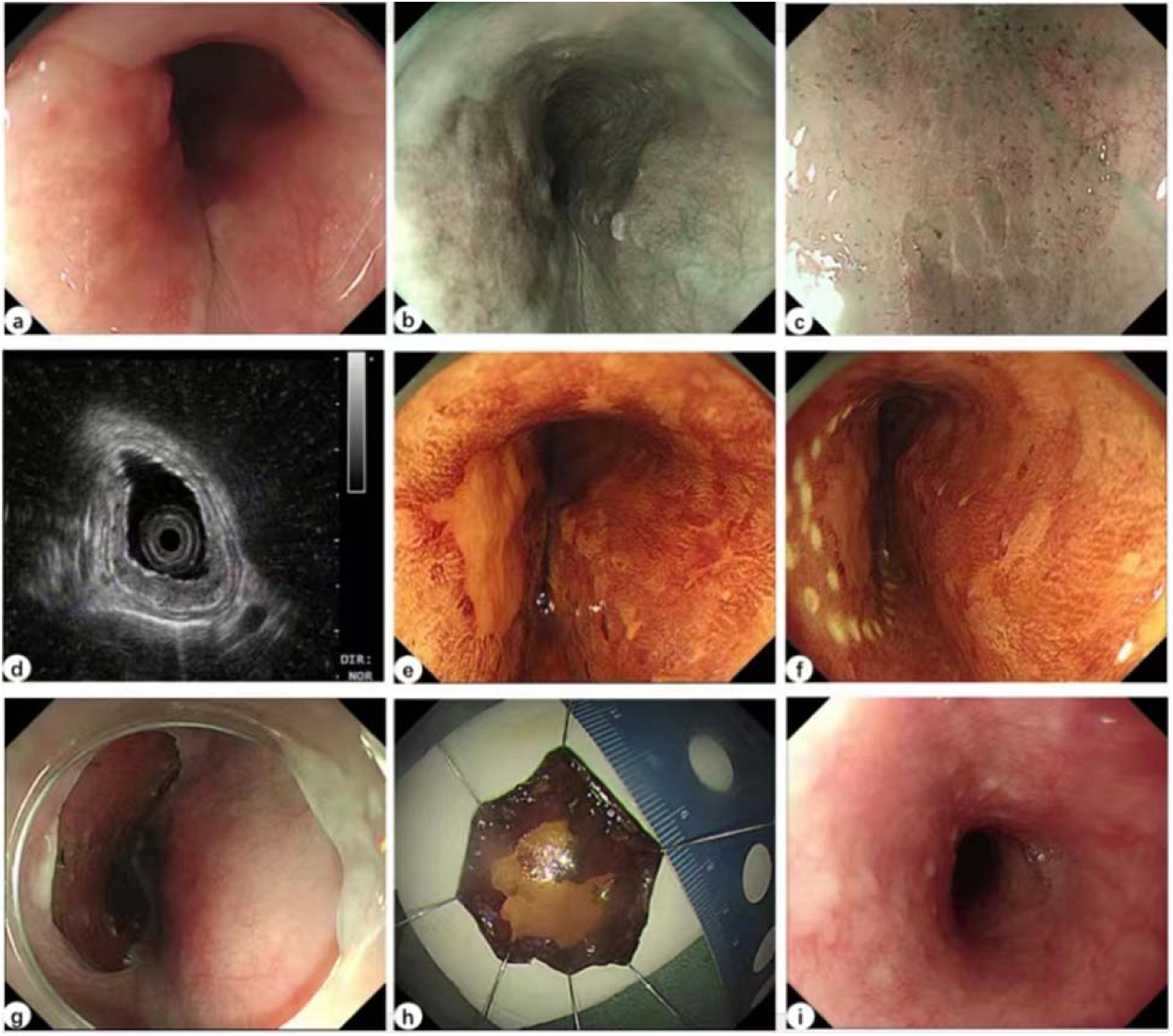
Representative images of an endoscopic mucosal dissection procedure for endoscopic resection of superficial esophageal squamous cell carcinoma. a. A white-light endoscopic image of an early lesion (0-IIb type), located 32-35 cm from the incisors, showed a rough, granular surface. b. Under narrow-band imaging endoscopy, the lesion was well-defined with a brownish color and occupied about 1/4 of the luminal circumference. c. The lesion was visible under magnified endoscopy with narrow-band imaging exhibited a B1 type intrapapillary capillary loop and small avascular area. d. With endoscopic ultrasonography, the lesion was hypoechoic with thickened mucosa and an intact submucosal layer. e. Lugol’s iodine staining demonstrated lamellar unstained mucosa and a positive pink sign for squamous cell carcinoma. f. Endoscopic lesion border marking with a DUAL knife was carried out about 0.3-0.5 cm away from the lesion edge with the aid of a transparent cap. g. After a complete endoscopic resection with endoscopic mucosal dissection, the lesion wound bed surface was smooth without bleeding, ready for a closure. h. The en blot fresh specimen was immediately pinned down flat for photography and subsequent fixation. i. Three months after the procedure, the lesion wound completely healed, demonstrated a smooth normal mucosal surface at the follow-up white-light upper endoscopy.

**Figure 3.**
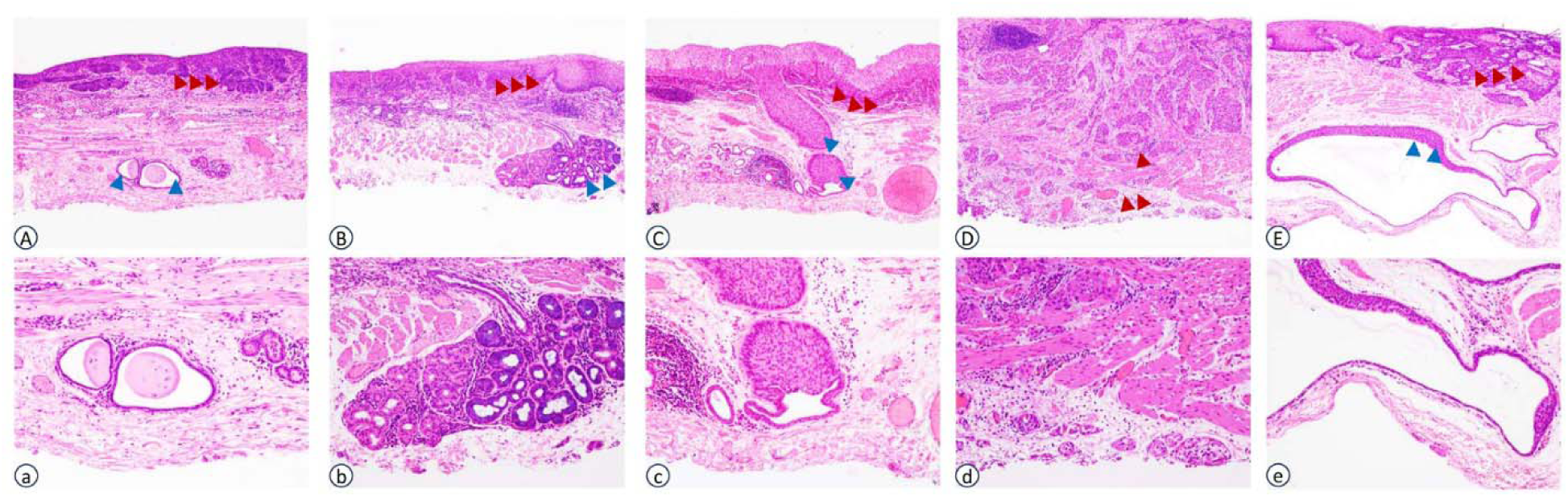
SESCC Histopathology. A-a. muscularis mucosae, NDI, vertical margin negative (HE ×40, HE ×100); B-b. submucosa, NDI, vertical margin negative (HE ×40, HE ×100); C-c. submucosa, DI, vertical margin negative (HE ×40, HE ×100); D-d. submucosa, NDI vertical margin positive (HE ×40, HE ×100); E-e. muscularis mucosae, DI, vertical margin negative (HE ×40, HE ×100). Carcinoma (red arrow), glandular ducts (blue arrow)

### Histopathological evaluation

The ESD-resected fresh tissue specimen was pinned down flat onto a dental vax board for photography and then immediately immersed in 10% formalin solution to be fixed overnight at the room temperature. All resection margins were inked with appropriate colors. Then, the specimen was sequentially cut into 2 mm-wide strips perpendicular to the base of a lesion, and embedded in paraffin for routine histopathologic evaluations. The histopathological diagnosis of SESCC followed the 5^th^ edition of the World Health Organization (WHO) criteria and the 16^th^ edition of the Japanese classification of esophageal cancer^5, 16^. The size of a tumor, the presence or absence of lymphovascular involvement (LVI), and the status of horizontal and vertical margins of an ESD resected specimen were recorded. The depth of tumor invasion was defined as follows: intra-epithelium (EP), lamina propria mucosa (LPM), muscularis mucosa (MM); superficial submucosa within 200 μm (SM1) or beyond 200 μm (SM2) below MM. *En bloc* resection referred to the ESD-resected tissue as a single piece. Pathologically, complete resection (R0) was defined as an *En bloc* resection with negative peripheral and vertical margins. Based on Japanese guidelines^5^, curative resection was defined as *en bloc* R0 resection for tumor invasion limited to the EP or LPM (pT1a-EP/LPM), well to moderately differentiated SESCC, and the absence of LVI in this study. However, because no recommendation has been made in the Japanese guidelines for SESCC with invasion confined to the MM (pT1a-MM) with negative LVI, those SESCC cases with LVI-negative pT1a-MM and pT1b-SM lesions were regarded with a potential risk of lymph node metastasis, and thus were considered as noncurative resections in this cohort^17^.

Based on the predominant histopathologic tumor growth and invasion front pattern (IFP) at the invasion front of SESCC, the following three patterns were classified: IFP-a (expansive), expansive growth of tumor nests with a well-demarcated border; IFP-b (intermediate), intermediate between IFP-a and IFP-c; and IFP-c (infiltrative), infiltrative growth of tumor nests and buds with an ill-defined border^5^.

### ESD safety assessment

In this cohort, the major intraoperative and perioperative ESD adverse events included immediate and delayed bleedings, perforation, and post-ESD stenosis. Immediate bleeding occurred intraoperatively and requires prompt hemostatic treatment, such as thermocoagulation and endoscopic clipping, while delayed bleeding, manifested by hematemesis or melena at 0–30 days after ESD, required emergency endoscopic interventions. Perforation was diagnosed when mediastinal connective tissue, such as fat, was observed intra-operatively, or the presence of air in the mediastinal space on plain chest radiography during an ESD procedure.

Esophageal stricture was defined as the presence of a markedly narrowed esophageal lumen, less than 1.0 cm in diameter, observed during post-ESD follow-up endoscopic evaluations, or the inability of a standard endoscope (with a diameter of approximately 1.0 cm) to passing through a previous ESD-resected esophageal lesion site. This condition was commonly associated with swallowing difficulties and required endoscopic dilation treatment^18, 19^.

### Post-ESD patient management and follow-up

Based on Hatta et al^17^, the SESCC tumors staged at pT1a-MM/pT1b-SM1 with negative LVI were regarded as non-curative because of potential LNM risk. Those patients were advised to undergo additional surgery, chemoradiation therapy, or close follow up^12^. Patients who achieved a curative resection were followed up endoscopically with or without biopsies at 3, 6, 12 months, and annually thereafter the ESD procedure. Enhanced chest computerized tomography was used to monitor the tumor occurrence, nodal/distant metastases, and scheduled at 6 and 12 months, and annually thereafter.

### Outcome data collection

Patient outcomes were assessed at regular clinical visits with a review of medical records and telephone interview of the patient or the family members until October 31, 2021. The 5-year overall survival (OS) was calculated from the date of the ESD procedure to the date of the last follow-up or death from any cause. The 5-year Disease-free survival (DSS) was calculated from the date of ESD to the date of death from esophageal squamous cell carcinoma. The number of dates was rounded up into month for statistical analysis.

### Statistical analysis

Differences in categorical variables were assessed with the Chi-square test or Fisher’s exact and reported as number and percentage, while differences in continuous variables were analyzed with the two-tailed Student’s t or the Mann-Whitney U test and tabulated as mean ± standard deviation (SD) or median with 25th-75th percentiles. The 5-year OS and DSS were estimated with the Kaplan–Meier method with a log rank test. A logistic regression model was used for univariate and multivariate analyses on risk factors of post-ESD stenosis. The results from the regression model were reported as odds ratio (OR) and 95% confidence interval (CI). SPSS 25.0 (IBM, Armonk, New York, USA) was used for all statistical analyses. A P value of < 0.05 was considered statistically significant.

## Results

### Demographic and clinical features

All 748 qualified ESD-treated SESCC patients were divided into either the DI (22.6%, 169/748) or NDI group (77.4%, 579/748). As shown in Table 1, there was a male predominancy with a male-to-female ratio of 1.6 and 2.1 for DI and NDI groups, respectively. The mean age of patients in the DI group was significantly older (average: 66.1 years, range 47-87), compared to those in the NDI group (mean: 64.5 years; range 36-88) (p < 0.05). There were no significant differences between the two groups in percentage of smoker, alcohol abuser, and most common comorbidities, such as hypertension, diabetes mellitus, reflux esophagitis, chronic atrophic gastritis, and family cancer history.

**Table 1.**
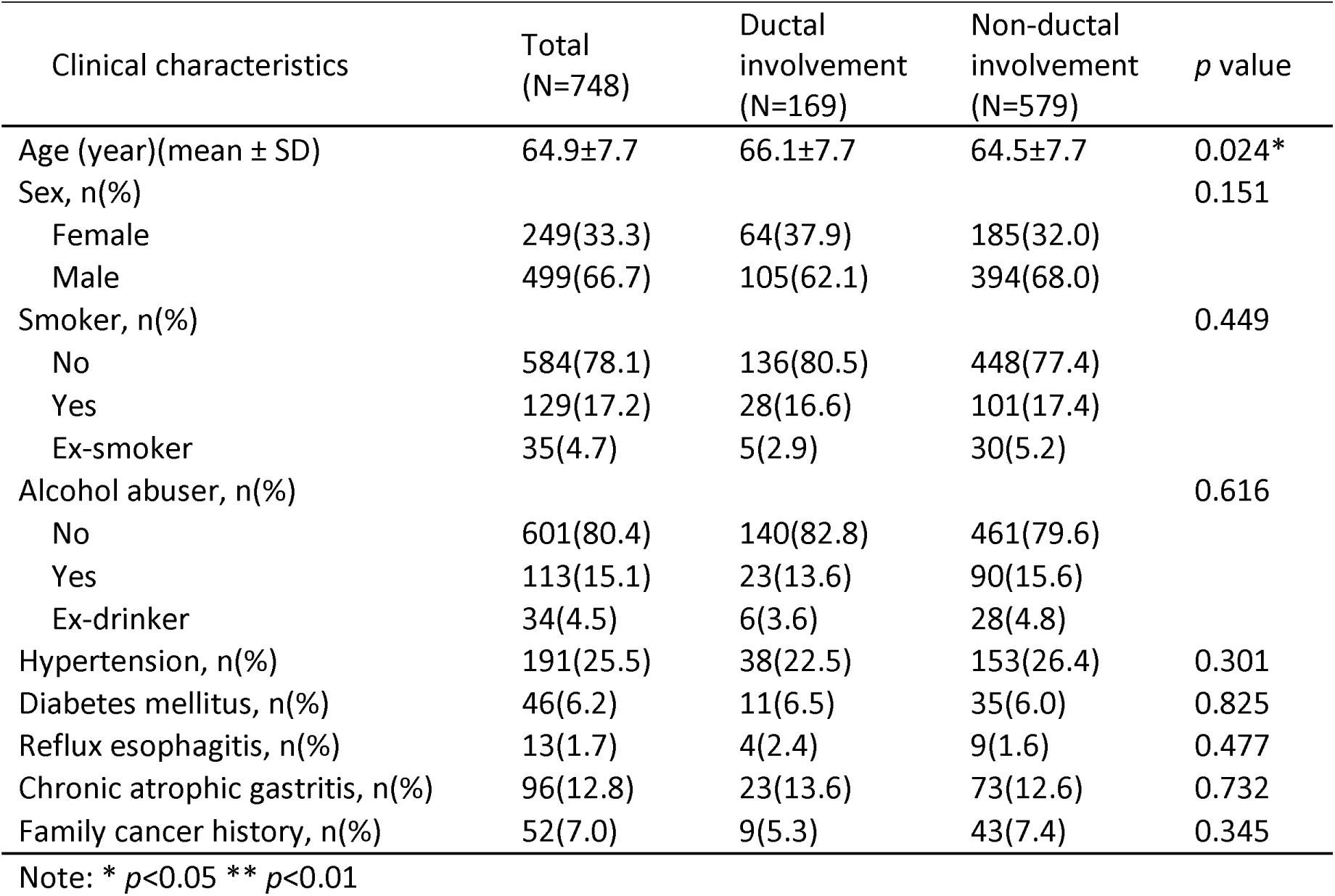
Demographic and clinical features of patients with superficial esophageal squamous cell carcinoma.

| Clinical characteristics | Total<br>(N=748) | Ductal<br>involvement<br>(N=169) | Non-ductal<br>involvement<br>(N=579) | p value |
| --- | --- | --- | --- | --- |
| Age (year)(mean ± SD) | 64.9±7.7 | 66.1±7.7 | 64.5±7.7 | 0.024* |
| Sex, n(%) |  |  |  | 0.151 |
| Female | 249(33.3) | 64(37.9) | 185(32.0) |  |
| Male | 499(66.7) | 105(62.1) | 394(68.0) |  |
| Smoker, n(%) |  |  |  | 0.449 |
| No | 584(78.1) | 136(80.5) | 448(77.4) |  |
| Yes | 129(17.2) | 28(16.6) | 101(17.4) |  |
| Ex-smoker | 35(4.7) | 5(2.9) | 30(5.2) |  |
| Alcohol abuser, n(%) |  |  |  | 0.616 |
| No | 601(80.4) | 140(82.8) | 461(79.6) |  |
| Yes | 113(15.1) | 23(13.6) | 90(15.6) |  |
| Ex-drinker | 34(4.5) | 6(3.6) | 28(4.8) |  |
| Hypertension, n(%) | 191(25.5) | 38(22.5) | 153(26.4) | 0.301 |
| Diabetes mellitus, n(%) | 46(6.2) | 11(6.5) | 35(6.0) | 0.825 |
| Reflux esophagitis, n(%) | 13(1.7) | 4(2.4) | 9(1.6) | 0.477 |
| Chronic atrophic gastritis, n(%) | 96(12.8) | 23(13.6) | 73(12.6) | 0.732 |
| Family cancer history, n(%) | 52(7.0) | 9(5.3) | 43(7.4) | 0.345 |

Note: \* $p < 0.05$ \*\* $p < 0.01$

### Endoscopic and pathological characteristics

As shown in Table 2, the most common location of SESCC was in the middle thoracic esophagus in the both groups with no significant difference among various locations. In contrast, the difference in macroscopic types between the two group was significant, in which 0-IIa, 0-IIb, and 0-IIc+IIa types were more frequently seen in the DI than in the NDI group (p < 0.01). The mean tumor size was 4.3 cm (range 1.4-8.7) for the cohort, and significantly larger in the DI (3.2 ± 1.8 cm) than in the NDI (2.6 ± 1.6 cm) groups (p < 0.01). On the other hand, the prevalence of endoscopic IPCL mucosal lesion types of B1+B2, B2, and B1/B2+B3 was significantly more common in the DI than in the NDI group (p < 0.01). Histopathologically, deeper tumor invasion and moderate/poor differentiations were significantly more frequent in the DI than NDI groups (p < 0.01 in both). The most common infiltration pattern at the tumor invasion front was IFP-a in both groups, but significantly more common in the DI (37.9%,64/169) than NDI (15.9%,92/579) groups (p < 0.01). The proportion of IFP-c was close in both groups, with 0.6% (1/169) in the DI group and 0.7% (4/579) in the NDI group.

**Table 2.**
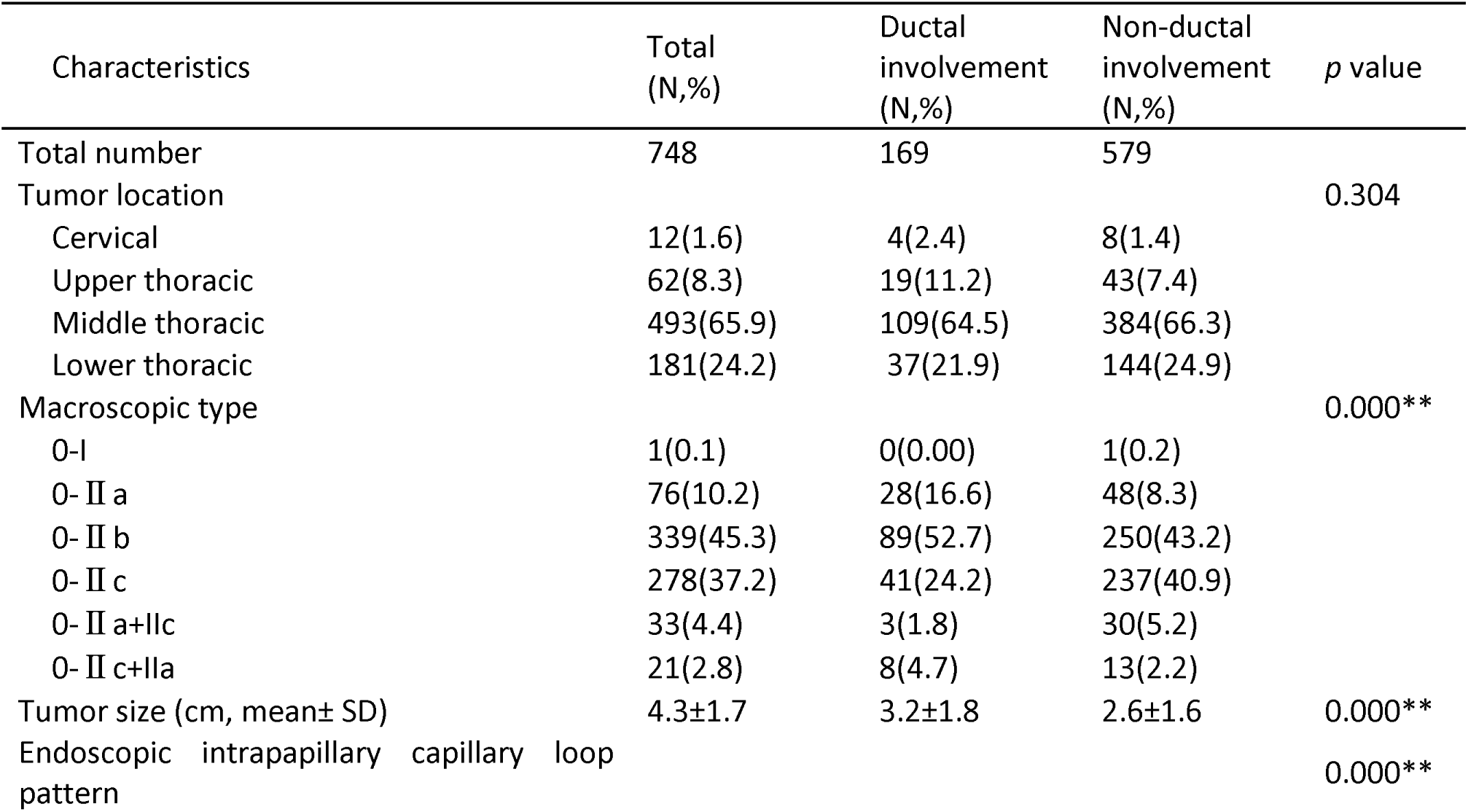
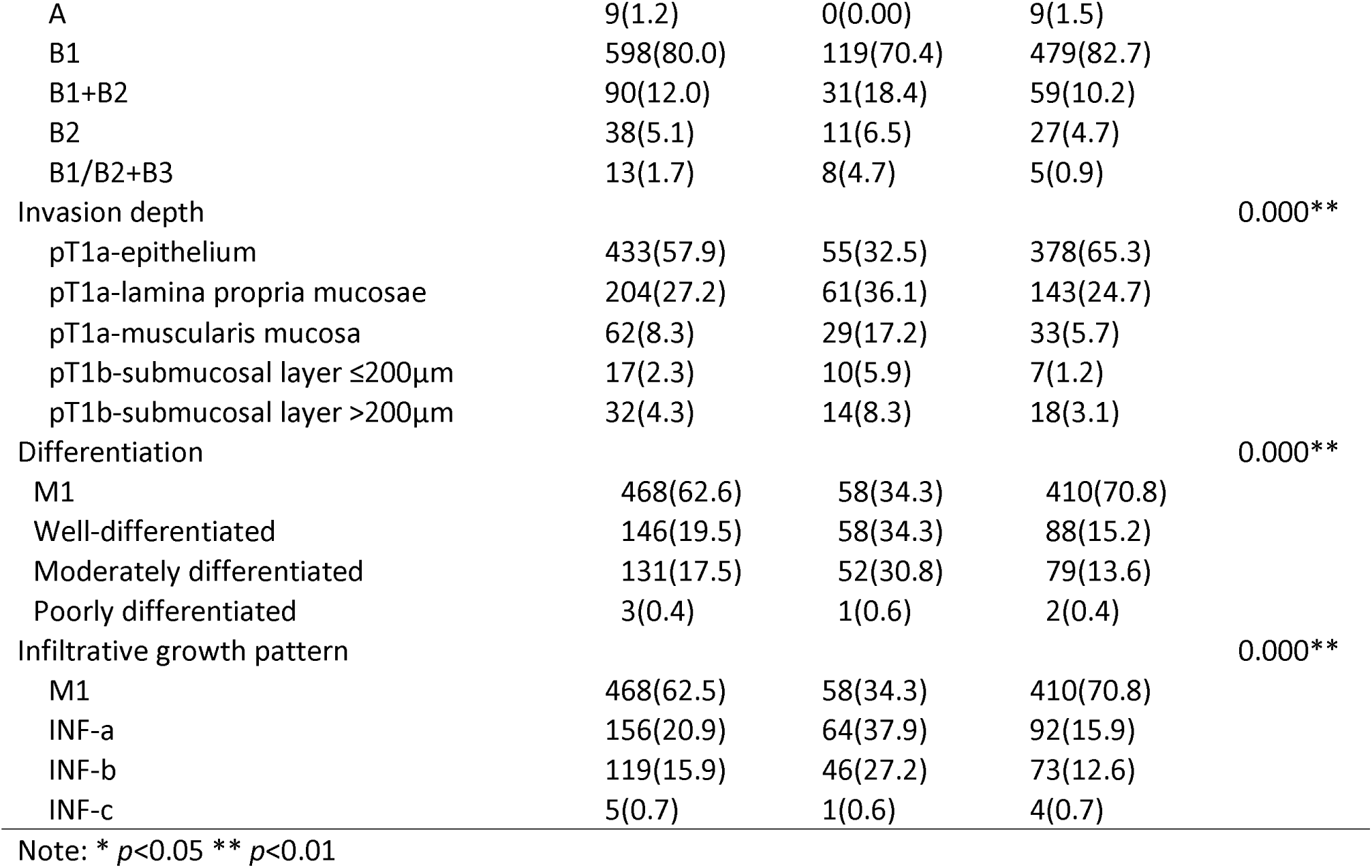
Endoscopic and pathological characteristics of superficial esophageal squamous cell carcinoma.

### ESD efficacy and safety

*En* bloc resection was 100% in both groups (Table 3). However, the complete resection rate was significantly lower in the DI than NDI (88.2% vs. 96.4%) groups (p < 0.01), and so as the curative resection rate (62.1% vs. 86.9%, p < 0.01). Non-curative resection occurred in 140 patients. The prevalence of a positive horizontal margin was significantly higher in the DI (10.1%, 17/169) than NDI (3.1%, 18/579) groups (p < 0.01), and so as the positive vertical margin (p < 0.05). There was no significant difference in lymphovascular invasion or hospital stay between the two groups (p > 0.05).Although there was no significant difference in overall ESD-related complications, including infection, bleeding, and perforation, between the two groups, postoperative esophageal stenosis occurred significantly more frequently in the DI than NDI groups (20.7% vs. 9.3%, p < 0.01).

**Table 3.** ESD efficacy and safety on resection of superficial esophageal squamous cell carcinoma.

| Efficacy and complication | Total<br>(N,%) | Ductal<br>involvement<br>(N,%) | Non-ductal<br>involvement<br>(N,%) | P value |
| --- | --- | --- | --- | --- |
| Total number | 748 | 169 | 579 |  |
| *En bloc* resection | 748(100.0) | 169(100.0) | 579(100.0) | 1 |
| Complete resection | 707(94.5) | 149(88.2) | 558(96.4) | 0.000** |
| Curative resection | 608(81.3) | 105(62.1) | 503(86.9) | 0.000** |
| Positive horizontal margin | 35(4.7) | 17(10.1) | 18(3.1) | 0.000** |
| Positive vertical margin | 9(1.2) | 5(3.0) | 4(0.7) | 0.017* |
| Lymphovascular invasion | 13(1.7) | 5(3.0) | 8(1.4) | 0.168 |
| Median hospital stay (day) (number, range) | 6(2-17) | 6(3-17) | 6(2-16) | 0.05 |
| Complication | 73(9.8) | 22(13.0) | 51(8.8) | 0.105 |
| Infection | 5(0.7) | 1(0.6) | 4(0.7) | 0.889 |
| Bleeding | 17(2.3) | 3(1.8) | 14(2.4) | 0.622 |
| Perforation | 1(0.1) | 0(0.0) | 1(0.2) | 0.589 |
| Stenosis | 89(11.9) | 35(20.7) | 54(9.3) | 0.000** |

\* $p<0.05$ \*\* $p<0.01$

### Risk factors for post-ESD esophageal stenosis

As shown in Table 4, with univariate analysis, statistically significant risk factors for post-ESD esophageal stenosis included male sex (p < 0.01), maximum diameter of the resected specimen > 5 cm (p < 0.01), circumferential extension of the tumor ≥ 3/4 (p < 0.01), middle or lower thoracic location (p < 0.05), DI (p< 0.01), and tumor infiltration up to SM2 (p < 0.05). In multivariate analysis, the independent risk factors for stenosis were male sex (OR: 0.53; 95% CI:0.32-0.87; p < 0.05), the maximal specimen size (OR:2.33; 95% CI:1.32-4.12; p < 0.01), circumferential extension ≥ 3/4 ( OR: 5.44; 95% CI: 2.97 – 9.97; p < 0.01), middle thoracic location (OR: 0.22; 95% CI: 0.06-0.87; p < 0.05), and lower thoracic location (OR: 0.19; 95% CI: 0.05-0.81; p < 0.05).

**Table 4.** Univariate and multivariate analyses of risk factors associated with post-ESD esophageal stenosis. Note: ESD:endoscopic submucosal dissection; SM: submucosa

| Variables | Post-ESD esophageal Stenosis |  |  |  |
| --- | --- | --- | --- | --- |
|  | Univariate | P | Multivariate | P |
|  | OR (95% CI) |  | OR (95% CI) |  |
| Age(years) |  |  |  |  |
| ≤65 | Ref. |  |  |  |
| > 65 | 1.25 (0.80-1.95) | 0.325 |  |  |
| Sex |  |  |  |  |
| female | Ref. |  |  |  |
| male | 0.44 (0.28-0.69) | <0.001 | 0.53 (0.32-0.87) | 0.013 |
| Maximum diameter of specimen(cm) |  |  |  |  |
| ≤5 | Ref. |  |  |  |
| >5 | 3.81 (2.39-6.07) | <0.001 | 2.33 (1.32-4.12) | 0.004 |
| Circumferential extension of the tumor |  |  |  |  |
| <3/4 | Ref. |  |  |  |
| ≥3/4 | 8.44 (5.02-14.18) | <0.001 | 5.44 (2.97-9.97) | 0.001 |
| Location |  |  |  |  |
| Cervical | Ref. |  |  |  |
| Upper thoracic | 0.76 (0.20-2.84) | 0.678 | 0.73 (0.17-3.09) | 0.664 |
| Middle thoracic | 0.24 (0.07-0.83) | 0.024 | 0.22 (0.06-0.87) | 0.031 |
| Lower thoracic | 0.18 (0.05-0.67) | 0.011 | 0.19 (0.05-0.81) | 0.024 |
| Ductal involvement | 2.54 (1.59-4.05) | 0.001 | 1.62 (0.92-2.83) | 0.092 |
| Depth of tumor invasion |  |  |  |  |
| M1 | Ref. |  |  |  |
| M2 | 1.04 (0.61-1.77) | 0.874 | 0.65 (0.35-1.19) | 0.162 |
| M3 | 1.39 (0.65-3.01) | 0.396 | 0.69 (0.28-1.71) | 0.422 |
| SM1 | 1.10 (0.24-4.94) | 0.906 | 0.78 (0.16-3.92) | 0.765 |
| SM2 | 2.74 (1.16-6.44) | 0.021 | 1.59 (0.58-4.35) | 0.362 |

Note: ESD: endoscopic submucosal dissection; SM: submucosa

### Long⍰term outcome

As illustrated in Table 5, the mean number of follow-up months was 44 (rang: 30-62) for the cohort and there was no significant difference between in the DI and NDI groups. During the follow-up period, additional surgical resection was carried out in 13 (1.7%, 13/748) patients for the cohort, 5(3.0%, 5/169) in the DI group and 8 (1.4%, 8/579) in the NDI group. Distant metastasis was found in one patient in the DI group and nine in the NDI group, but the difference was not significant between the two groups. Chemoradiation therapy was needed in 13 (1.7%) patients in the cohort and there was no significant difference between the two groups (2.4% in DI and 1.6% in NDI groups). For a variety of reasons, 23 patients in the NDI group did not undergo additional surgery as recommended. Three patients in the NDI group of those declined additional therapy died of the disease. One patient in the NDI group who underwent additional surgery died of the disease. The 5-year OS rate was 95.7% for the cohort and significantly higher in the DI (100%) than NDI (94.6%) (p < 0.05, Log-Rank Test). The 5-year DSS rate was 93.9% for the cohort and the difference was not significant between the two groups (Table 5, Figure 4 and Figure 5)

**Figure 4.**
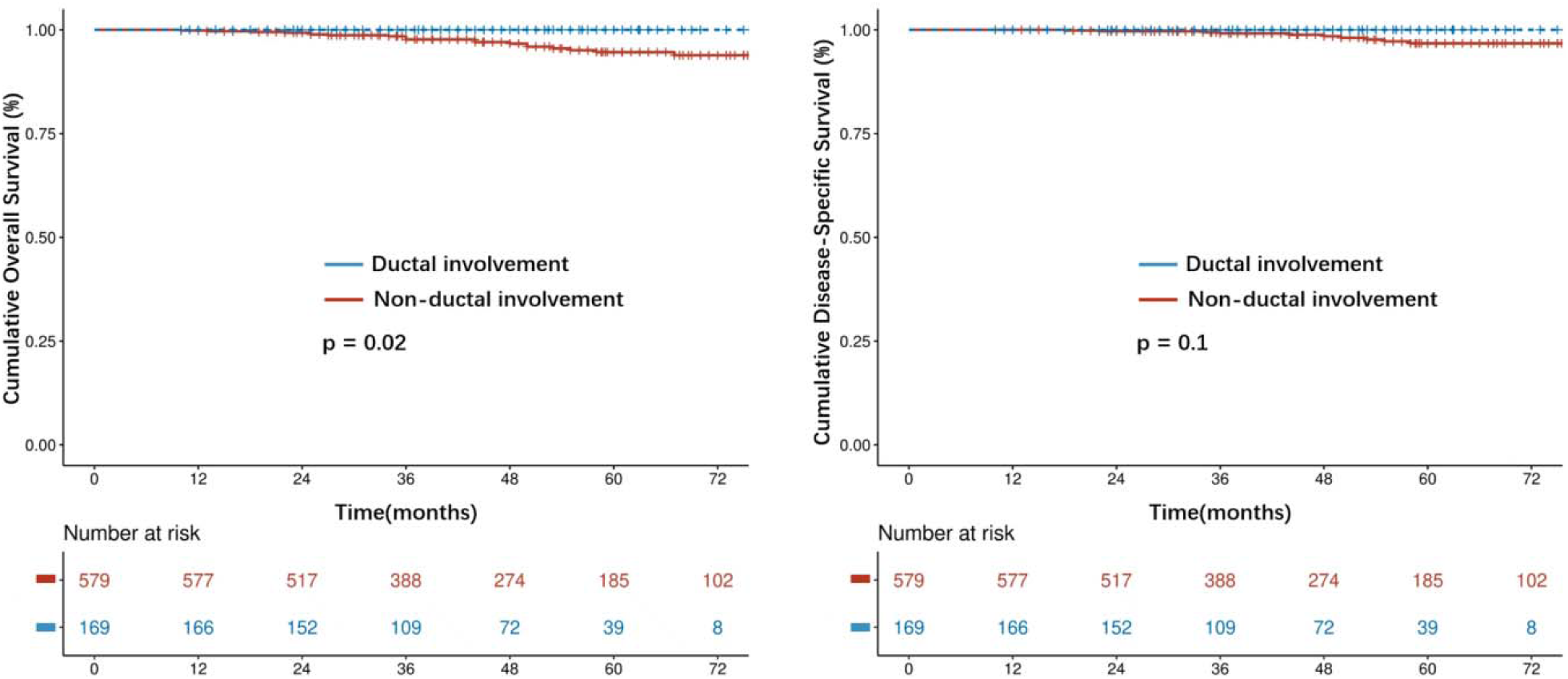
Kaplan-Meier curves in patients with and without ductal involvement. (A) Overall survival. (B) Disease-specific survival.

**Figure 5.**
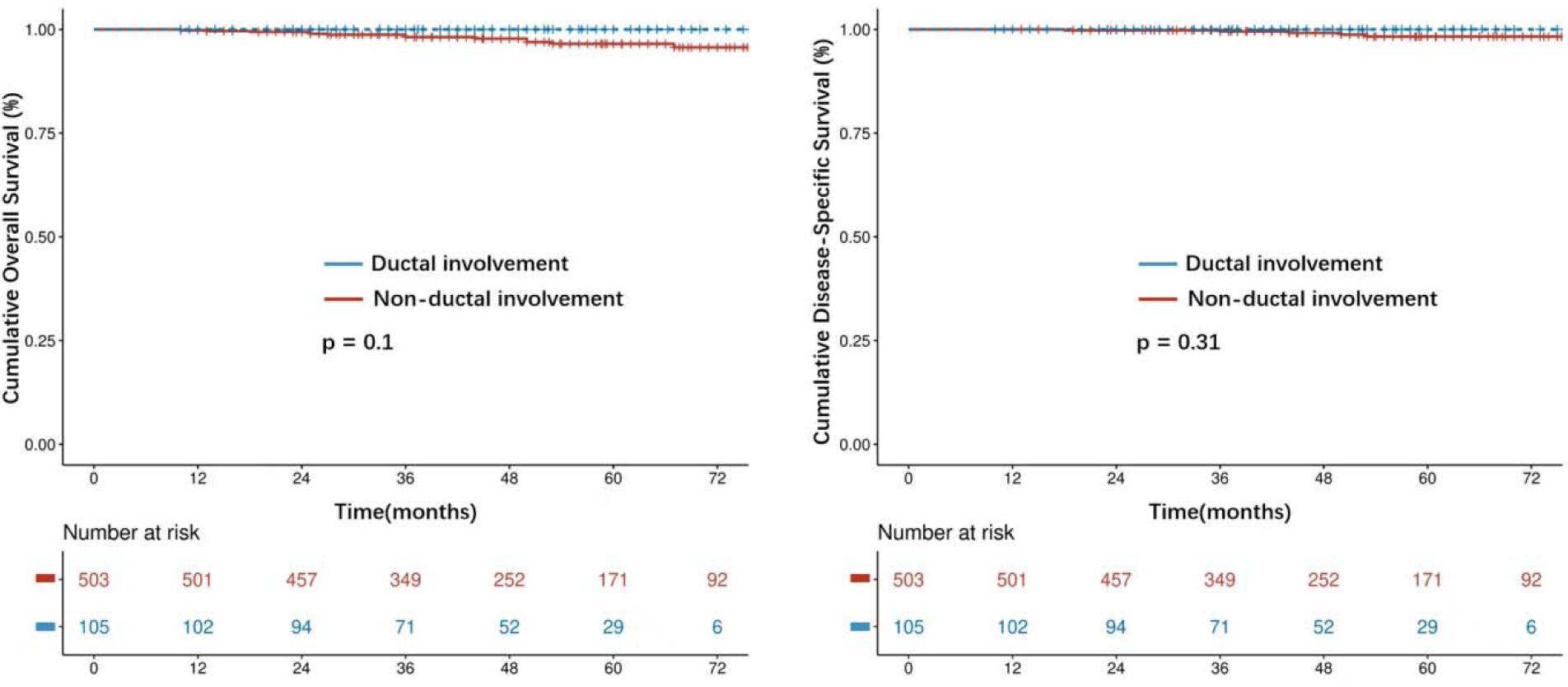
Kaplan-Meier curves for all curatively resected patients with and without ductal involvement. (A) Overall survival. (B) Disease-specific survival.

**Table 5.** Comparison of post-endoscopic dissection outcomes between SESCC patients with or without ductal involvement.

| Outcome | Total<br>(N,%) | Ductal<br>involvement<br>(N,%) | Non-ductal<br>involvement<br>(N,%) | *p* value |
| --- | --- | --- | --- | --- |
| Total number | 748 | 169 | 579 |  |
| Follow-up (month), median (25%,75%) | 44(30.0,62.0) | 43(29.0,59.0) | 45(30.0,64.0) | 0.058 |
| Additional Surgery | 13(1.7) | 5(3.0) | 8(1.4) | 0.168 |
| Distant metastasis | 10(1.3) | 1(0.6) | 9(1.6) | 0.338 |
| Chemoradiotherapy | 13(1.7) | 4(2.4) | 9(1.6) | 0.478 |
| 5-year Overall survival rate | 712(95.7) | 169(100.0) | 548(94.6) | 0.020* |
| 5-year Disease-specific death | 729(97.4) | 169(100.0) | 560(96.7) | 0.102 |
| Recurrence-free survival | 702(93.9) | 163(96.5) | 539(93.1) | 0.11 |

Note: SESCO: superficial esophageal squamous cell carcinoma

## Discussion

In this single-center retrospective study on ESD-resected SESCC with (DI) or without (NDI) the involvement of esophageal gland ducts, our results revealed SESCC-DI tumors with a significantly larger tumor size, deeper invasion, lower complete and curative resection rates, and more frequent post-ESD stenosis, compared to SESCC NDI tumors; however, there were no significant differences between the two groups in ESD-related complications, disease-specific death, recurrence-free survival, distant metastasis, and the requirement for subsequent chemoradiation therapy and/or surgery. Therefore, our data, if confirmed in the future studies with larger samples, suggest that ESD was safe with outstanding efficacy, minimal complications, and excellent prognosis for endoscopic resection of SESCC with or without esophageal gland duct involvement.

DI is not uncommon in esophageal squamous cell carcinoma, and its incidence has been reported in the literature to be 11.6%-38.1%^7, 14^. The presence of DI may influence the therapeutic decision-making process, especially when tumor cells reach the submucosal layer along the esophageal glandular ducts, which may lead to tumor recurrence and worse long-term survival^13, 20^. In a study that included 30 cases of early esophageal squamous cell neoplasia successfully treated with RFA, Wang *et al*. found that 7 local recurrences occurred in 6 patients during follow-up, and histopathology showed that DI was present in 6/7(85.7%) lesions and all were deeper than the muscularis mucosae, which may have contributed to the recurrences, whereas RFA can be ablated up to a maximum depth of the muscularis mucosae. A prospective study that included 210 patients with SESCC suggested that DI was an indicator of poorer prognosis (p <0.05), and among 185 patients with SESCC who met the criteria for curative resection of ESD (R0 resection and absence of deep submucosal infiltration or LVI), patients with DI had a lower rate of recurrence-free survival than patients in the NDI group (log-rank p < 0.05), and a local recurrence risk was higher (12.7% vs. 2.5%, HR=4.20, p < 0.05)^13^. However, Chen^21^ et al. strongly supported ESD treatment for SESCC. Prof. Chen emphasized the complete resection (including submucosa) in ESD treatment of superficial esophageal cancer, with vertical resection as close as possible to the surface of the muscularis propria and covering the entire submucosa as much as possible, in order to completely resect the ducts that may be involved.

Before the advent of endoscopic techniques, esophagectomy(ESO) was the treatment of choice for SESCC. Tadakazu and colleagues studied 201 surgically resected lesions in 140 SESCC patients and found no significant differences in DI with respect to LVI, LNM, and prognosis. They concluded that DI is not significant as a pathway for tumor spread in esophageal squamous cell carcinoma following surgical resection^7^.The use of endoscopic treatment for stage T1 esophageal cancer has increased significantly in recent years years^22^ The goal of endoscopic treatment is to provide potentially curative options for patients with superficial cancers while preserving the integrity of the esophagus. Several recent studies have emphasized the preference for ER over radical surgery in appropriately selected patients with superficial esophageal cancer^23–25^. A meta-analysis of 16 studies suggested that ESD-treated SESCC had a shorter hospital stay (Hedge’s g: –1.22) and shorter operative time (g: –4.54) than ESO, with a significant reduction in the risk of ESD complications (OR: 0.35) and all-cause mortality (OR: 0.56). There were no significant differences in recurrence rate (OR: 0.95), short-term outcome (OR: 1.10) and long-term survival (OR: 0.81) between ESD and ESO^26^. Additionally, the ESD technique has been continuously advancing. Takahashi et al. reported a case of SESCC-DI treated with underwater endoscopic submucosal dissection(UESD) under general anesthesia. This updated ESD technique can prevent positive vertical margins in SESCC-DI^27^.

Patients in the DI group had a poorer short-term prognosis, with lower rates of complete and curative resection than those in the NDI group. In addition, SESCC-DI patients who underwent ESD had a higher probability of developing stenosis (p<0.01), with about 1/5 patients in the DI group and about 1/10 patients in the NDI group. One of the common complications after ESD is esophageal stricture, which can lead to severe dysphagia in the patients, thus greatly reducing their quality of life (QOL)^28^. In addition, frequent and prolonged endoscopic balloon dilatation (EBD) can be financially and psychologically stressful for patients^29^ Currently, prophylactic EBD, steroid injection therapy, and oral steroid therapy can prevent esophageal strictures^29,30^. Frequent endoscopic balloon dilatation (EBD) can be performed to treat benign esophageal strictures, but it may also lead to serious complications such as perforation and other adverse events^31^ Past studies have pointed out that during ESD for SESCC, the margins of vertical resection should be close to the surface of the muscularis propria to encompass as much as possible all the submucosal layers so as to completely resect possible infiltrating lesions for curative resection^13, 21^ However, we should be mindful of the need for more experienced endoscopists to perform ESD in SESCC-DI patients. Chen commented that SESCC should be performed with care and skill when performing ESD to avoid major complications^21^ The endoscopist needs to perform delicate maneuvers in order to achieve curative resection and reduce esophageal stricture while avoiding damage to the esophageal muscle. If ESD is used for SESCC-DI, we should obtain informed consent from patients to inform them of the possible risk of esophageal stricture. However, Tang et al. found that an independent risk factor for refractory esophageal stricture after extensive ESD was a resection length of ≥50 mm, and the risk of esophageal stricture would be significantly increased if the resection length was ≥50 mm^32^ Our study also had larger tumors in the DI group, which may have also influenced the occurrence of strictures after ESD in this group. In the risk factor analysis for postoperative stricture, we found that DI was not significant in the multifactorial analysis, probably because of its covariation with the size of the resected specimen and gender, but the lower limit of the 95% CI of its OR in the multifactorial analysis was close to 1, which may also be a risk factor for postoperative stricture in ESD. The effect of DI on the occurrence of stricture after ESD may need to be further confirmed in a large sample study. This study also analyzed a subgroup of endoscopically presented type B1 vessels only. According to IPCL typing, most of the B1 vessels found endoscopically were SESCC with infiltration-guided EP/LPM. our study demonstrated that a larger proportion of submucosal infiltration occurred in the DI group (p<0.01). According to the guidelines (ER Guidelines for Esophageal Cancer in Japan, 2020), SESCC with pT1b-SM requires additional surgery or radiotherapy because of the high risk of LNM. If we assess the depth of infiltration of the lesion only by the B1 vessels observed endoscopically, then the endoscopist may appear to underestimate the depth of invasion, but if it enables radical resection, the patient’s survival may not be affected.

Our study demonstrated that there was no significant difference in the long-term prognosis between the two groups after curative resection (p > 0.05). In 1993, Tadashi et al. concluded that even intramucosal esophageal cancer may have intraductal spread and that ER should be able to completely resect the residual tumor portion of submucosal esophageal glands^33^.ESD is an endoscopic resection technique that allows deeper penetration into the muscular layer of the mucosa^34, 35^. This may explain the lack of statistical difference in DSS between the DI and NDI groups of patients. In addition, previous studies have shown that tumor infiltration pattern (INF) is an independent factor influencing tumor prognosis and a significant predictor of prognosis in esophageal, gastric, and colorectal cancers^36–39^.IFP-c is an independent risk factor for lymph node metastasis and an independent prognostic factor for poor prognosis in stage T1 esophageal squamous cell carcinoma (ESCC)^39^. In our study, the proportion of INF-c was similar in both groups, which may be related to the fact that there was no statistically significant difference in the long-term prognosis between the two groups. Wang et al. found in a retrospective study of 210 patients with SESCC who underwent ESD surgery that the presence of DI was associated with poorer long-term survival and higher risk of local recurrence after curative resection with ESD^13^. This is contrary to the results of the present study, but the sample size of the present study was much larger, and more in-depth prospective studies are needed in the future to validate this result.

There are several limitations to this study. Firstly, this is a single-center study. Due to the fact that only patients who received treatment at the center were included in the study, the sample selection is limited and susceptible to selection bias. As a consequence, the findings may not be presentative of the entire patient population. However, this study encompassed the highest proportion of patients with SESCC undergoing ESD who satisfied the predefined inclusion criteria for analysis. Secondly, retrospective studies are susceptible to confounding factors. To reduce the impact, we reviewed the medical records of patients one by one, and used strict inclusion and exclusion criteria to ensure that the study and control groups were as similar as possible, which increased the reliability of the research results. Thirdly, this is data from a tertiary care endoscopy center in Jiangsu province, where ESD procedures are performed by experienced endoscopists, but the results may not apply even to the group of patients in all centers. This study is a single-center data, which still needs to be further verified with multi-center and prospective data.

In conclusion, our study shows that ESD is a successful and relatively safe treatment for SESCC with DI, especially for experienced endoscopic teams. Compared with NDI group, ESD is more suitable for DI lesions with shallow tumor invasion depth and small area, and attention should be paid to prevent postoperative stenosis. However, multicenter studies are needed to further verify our data.

## Author Disclosure Section

Yanan Wang, Zhiwen Li, Hai Wu, Xinlu Zhao, Shangtao Mao, Zhenyu Wang, Ying Yuan, Qiange Ye, Yanmei Zhu, Ying Xiang, Qin Huang, Lei Wang, Guifang Xu have no conflicts of interest or financial ties to disclose.

